# Etiologies and sequelae of extreme thrombocytosis in a large pediatric hospital

**DOI:** 10.1101/2020.06.01.20119438

**Authors:** Christopher S Thom, Emily Echevarria, Ashley D Osborne, Leah Carr, Kathryn Rubey, Elizabeth Salazar, Danielle Callaway, Thomas Pawlowski, Matt Devine, Stacey Kleinman, John Flibotte, Michele P Lambert

**Author notes:** Corresponding Author: Christopher S Thom Children’s Hospital of Philadelphia 3401 Civic Center Blvd 2NW46, Division of Neonatology Philadelphia, PA 19146.

## Abstract

Extreme thrombocytosis (ET, platelet count >1000 × 10^3^/ul) is an uncommon clinical finding 1. Primary ET is associated with myeloproliferative disorders, such as essential thrombocythemia 2. Secondary ET is more common and occurs in reaction to infection, inflammation, or iron deficiency. Bleeding and thrombotic complications more frequently arise in primary ET cases 1, but have been reported with secondary ET in adults 3. Etiologies and complications associated with ET in children are less well-defined, as prior pediatric studies have been relatively small or restricted to specialized patient populations 4,5. We aimed to characterize ET in a large, single-center pediatric cohort.

## Main Text

Extreme thrombocytosis (ET, platelet count >1000 × 10^3^/µl) is an uncommon clinical finding ^1^. Primary ET is associated with myeloproliferative disorders, such as essential thrombocythemia ^2^. Secondary ET is more common and occurs in reaction to infection, inflammation, or iron deficiency. Bleeding and thrombotic complications more frequently arise in primary ET cases ^1^, but have been reported with secondary ET in adults ^3^. Etiologies and complications associated with ET in children are less well-defined, as prior pediatric studies have been relatively small or restricted to specialized patient populations ^4,5^. We aimed to characterize ET in a large, singlecenter pediatric cohort.

### Patient identification

Utilizing the electronic medical record, we identified patients with ET who were admitted to our quaternary pediatric hospital from 2012 to March 2020. We retrospectively determined ET etiologies in these patients, along with associated ICD-9 diagnoses, treatments, and complications. Each hospital admission in which a patient had ET was considered an independent case. Most had multiple blood counts showing ET-range platelet values, giving us confidence that these findings did not result from laboratory error. Patient ages were assigned based on the first ET-range platelet count value in each admission. This study was deemed exempt from review by the Children’s Hospital of Philadelphia Institutional Review Board.

We defined one principal etiology for each ET case. ‘Infection’ cases included those with culture-proven bacterial infections, laboratory-confirmed viral infections, and patients who received antimicrobials for suspected infection. Inflammation cases included those with clinical or laboratory evidence of inflammatory processes without confirmed infection (e.g., elevated C-reactive peptide values), as well as instances where malignancy, chemotherapy, trauma, or post-surgical changes were deemed principal causes. Iron deficiency cases were identified by ICD-9 diagnosis codes and/or supplemental iron prescriptions, in the absence of infection or signs of inflammation. ‘Other’ cases were those without a clear inciting cause. Many cases were potentially multifactorial, but typically involved one change (e.g., new infection) that was assigned as the principal etiology on review.

### Etiologies and sequelae of extreme thrombocytosis in a pediatric population

Of 79,618 patients in 131,982 admissions evaluated, we identified ET in 392 patients (0.5%) during 425 hospital admissions (0.3%, **Table 1**). For comparison, 15% of all patients admitted to our hospital had thrombocytosis >500 × 10^3^/µl over the same time period. At least 92% of our ET cases were secondary to infection (47%), inflammation (40%), or iron deficiency anemia (5%, **Fig. 1A**). Other etiologies accounted for 8% of cases, including bone marrow reactivity after hemorrhage (n=2), cases with no clear cause, and 1 potential case of primary ET in the setting of a novel genetic mutation ^6^. Overall, this distribution of etiologies in our cohort was consistent with prior pediatric ET reports ^4,5^.

**Table 1.** Characteristics of patients analyzed for this study. A total of 79,618 patients and 131,982 admissions were queried to identify patients meeting inclusion criteria. Temporal associations were defined as ±2 weeks from the first laboratory identification of ET.

| Characteristic | Value |
| --- | --- |
| Patients included (% of all patients) | 390 (0.5%) |
| Male patients included (%) | 195 (50%) |
| Patient age at time of ET (years, mean±SD) | 5.2 ± 6.0 |
| Age range at time of ET | 1 d – 25 yrs |
| Admissions included (% of all admissions) | 423 (0.3%) |
| Principal etiology: |  |
| Secondary/reactive thrombocytosis | 422 |
| Primary thrombocytosis (suspected) | 1 |
| Intensive care unit admissions (%) | 231 (55%) |
| Temporally associated thrombotic event (%) | 21 (5%) |
| Occurred after ET-range platelet count | 1 |
| Temporally associated bleeding event (%) | 6 (1%) |
| Occurred after ET-range platelet count | 0 |
| Empiric aspirin for ET (%) | 25 (6%) |

**Figure 1.**
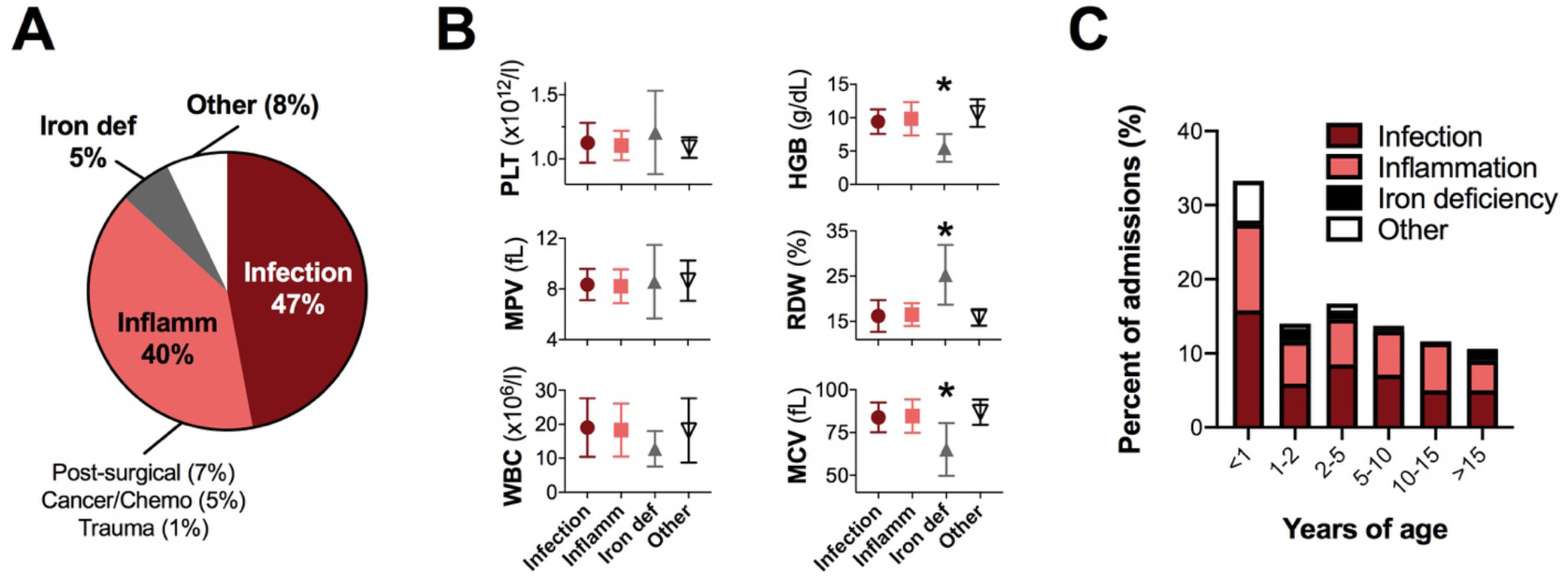
Etiologies, blood parameters and ages of patients with extreme thrombocytosis. **A**. Etiologies of ET in our cohort. Cases were assigned to categories based on principal factors underlying ET (infection, inflammation, iron deficiency anemia), or ‘other’ if there was no principal cause identified. **B**. Hematologic indices for cases in each category (mean±standard deviation). *p< 0.05 by ANOVA versus all other categories. **C**. Ages at which patients had extreme thrombocytosis. For each case, the first blood count showing ET was used to calculate patient age. Colors within each bar depict etiology category breakdown within each age range.

Hematologic indices showed marked microcytic anemia in the context of iron deficiency cases, and leukocytosis in cases of infection or inflammation (**Fig. 1B** and **Supplemental Table 1**). Indices in ‘other’ cases suggested robust multilineage hematopoiesis, with high red cell distribution width (RDW) and mean platelet volume (MPV) values indicating the presence of reticulocytes and immature platelets, but did not significantly differ from infection or inflammation cases. This could indicate hyperactive bone marrow following myelosuppression ^1^, which we viewed as a diagnosis of exclusion for our study.

Pediatric ET is generally not associated with bleeding or thrombotic sequelae ^4,5^. Despite this, aspirin was initiated for 25 patients in our cohort in response to laboratory values alone. This likely underestimates use of this therapy, since children with non-extreme thrombocytosis outside of the scope of this analysis may have also been treated empirically. There were 6 bleeding and 21 thrombotic events diagnosed within ±2 weeks of ET development. In all but one case, these events preceded ET onset, suggesting they were inciting factors as opposed to sequelae. For one patient, ET preceded by 1 day a central line-associated thrombus. However, this child was medically complex, in acute respiratory failure from rhinovirus and *E. coli* sepsis, and was already taking aspirin for thrombocytosis. Therefore, it is impossible to establish causality or definitive attribution to ET. Overall, our findings confirmed that bleeding or thrombosis from ET are incredibly rare in the pediatric population.

### Young age and critical illness correlate with extreme thrombocytosis

Infants and young children were overrepresented in our cohort, with 33% of patients less than 1 year old and almost half (47%) under 2 years old (**Fig. 1C**). By comparison, 30% of all admitted patients were under 2 years old in the queried time period (p=0.02 by two-sided Fisher’s exact test).

Interestingly, most ET cases without clear etiology occurred in infants less than 1 year old (p= 0.003 by two-sided Fisher’s exact test, **Fig. 1C**). It is possible that ET occurred more frequently in infants whose immature hematopoietic systems responded differently to pathophysiologic processes than older children. The hyperproliferative capacity of neonatal megakaryocytes, relative to adult megakaryocytes ^7^, may also underlie the preponderance of infants in our cohort.

Exaggerated pathophysiology during critical illness might predispose to more extreme hematologic derangements. Indeed, 55% of our cohort required support in our pediatric (PICU), neonatal (NICU), and/or cardiac intensive care units. Central venous access, an iatrogenic thrombosis risk factor^8^ and indication of critical illness, was in place for 29% of our cases.

Most critically ill children with ET were admitted to the PICU (73%), with a distribution of etiologies reflective of our general patient population (**Fig. 2A**). This proportion of PICU patients with ET (0.6%) was less than a prior report (∼1.1%)^4^. A further 21% of critically ill patients with ET were cared for in the NICU, which corresponded to 0.5% of all NICU patients in the queried time period.

**Figure 2.**
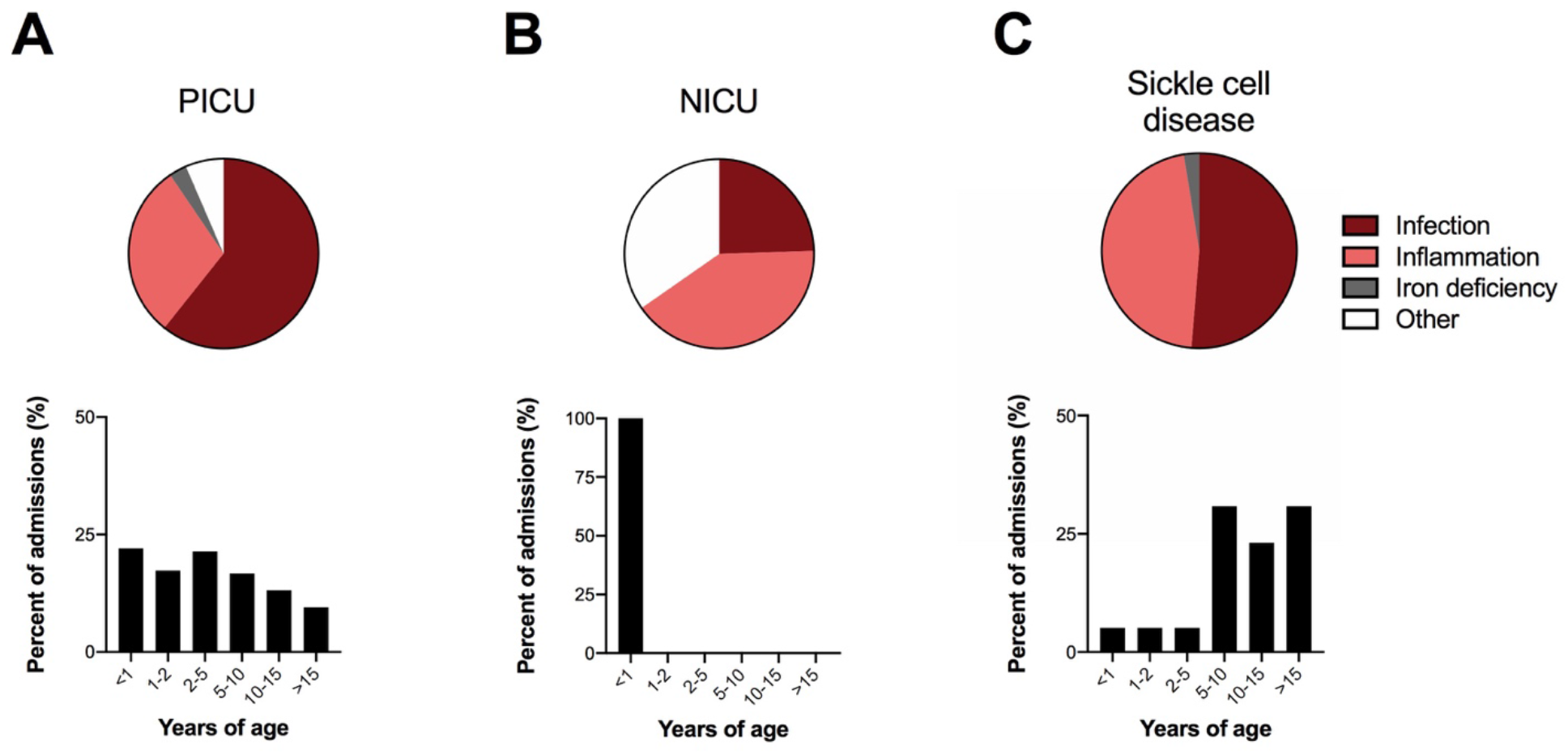
Etiologies, ages and blood parameters for exemplary patients with ET, including those hospitalized in the PICU, NICU, or with sickle cell anemia. **A**. Pie chart shows principal etiologies for ET in patients hospitalized in the PICU. Bar plot shows ages of PICU patients with ET from any cause. **B**. Pie chart shows principal etiologies for ET in patients hospitalized in the NICU. Bar plot shows ages of NICU patients with ET from any cause. **C**. Pie chart shows principal etiologies for ET in patients with sickle cell disease. Bar plot shows ages of sickle cell disease patients with ET from any cause.

Remarkably, 35% of NICU patients had an unclear etiology for ET (p< 0.001 by two-sided Fisher’s exact test vs. all other ICU patients, **Fig. 2B**). In these patients, ET frequently occurred just prior to discharge, without related clinical changes. We suspect that all or most of these cases represented hyperactive bone marrow following myelosuppression due to critical illness, as suggested by robust multilineage hematopoiesis in these infants (**Supplemental Table 2**).Blood count indices did not vary between etiological categories in these age-matched patients.

### Diagnoses associated with extreme thrombocytosis

Respiratory infections have been linked to ET ^5,9,10^ and were common in our cohort. Bacterial and viral lung infections were diagnosed in 49% of infection-associated ET cases. Although infants and young children may be at higher risk for hospitalization from respiratory infections (e.g., bronchiolitis), lung infections were not preferentially diagnosed in young patients within our cohort (52% of children < 2 years old vs. 48% of children > 5 years old with ET from infection). Thus, respiratory infections did not account for the prevalence of infants in our cohort.

Asplenia can cause thrombocytosis due to reduced platelet storage and removal ^5^. Sickle cell disease leads to autosplenectomy and functional asplenia by ∼5 years of age ^11^. In our cohort, 32 patients with sickle cell disease had ET during 39 hospitalizations. This was somewhat lower than expected, given our large sickle cell program and their predisposition to encapsulated bacterial infections. Principal ET etiologies for sickle cell patients virtually always involved vasoocclusive sickle cell crises, with or without infection (**Fig. 2C**). The age distribution for sickle cell patients presenting with ET was consistent with functional asplenia onset, as 85% occurred after age 5 (**Fig. 2C**). These exemplary cases show principal events (e.g. infections) driving ET only in a permissive context (e.g., underlying asplenia).

Thrombocytosis can occur in the setting of inflammatory diseases (e.g.,^12–15^). Patients with inflammatory conditions were frequently identified in our cohort, including 24 patients with inflammatory bowel disease (IBD) ^12^, 14 patients with Kawasaki disease ^13^, 5 patients with nephrotic syndrome ^14^, and 2 patients with juvenile idiopathic arthritis ^15^. The relatively high number of patients with IBD may be due to disease prevalence, iatrogenic thrombocytosis from steroid use, and/or the potent thrombocytosis-inducing effects of cytokines produced during intestinal inflammation ^12^.

### Genetic variation may contribute to developing extreme thrombocytosis

Our results suggested that individual genetic variation may contribute to ET, in addition to clinical and environmental factors. It was unclear why certain individuals developed ET, while others with identical conditions did not. For example, the reasons that a particular subset of 32 patients with sickle cell disease developed ET were unknown. We also suspected underlying genetic contribution in 61 patients who had recurrent ET (³2 independent episodes). Some of these blood counts occurred as outpatients, so were not included in our primary analysis.

In sum, our findings confirm common etiologies and a paucity of clinical sequelae related to ET in pediatric patients. These results may help inform clinical decision-making regarding empiric treatment strategies. Our cohort included many infants, several of whom had unexplained ET. These cases may reflect hyperactive bone marrow hematopoiesis following myelosuppression, and/or differential effects related to neonatal hematopoiesis. Our results also suggest that genetic variation could predispose some individuals to ET. Future studies to better understand such genetic factors may lead to novel insights into hematopoiesis, megakaryopoiesis, and/or thrombopoiesis.

## Data Availability

De-identified summary data will be made available upon request.

## Acknowledgements

The authors thank the patients and families for whom we are privileged to care at the Children’s Hospital of Philadelphia. We are grateful for incredible support from Scott Lorch, Heather French and the Children’s Hospital of Philadelphia Neonatal and Perinatal Medicine Fellowship program. We thank Russell Kesman and Kuan-Chi Lai for their thoughtful suggestions.

## Contributions

CST designed the study. All authors analyzed and interpreted data. CST wrote the paper. All authors reviewed, revised and approved the final version of the manuscript.

